# A 2-year calorie restriction intervention reduces glycomic biological age biomarkers

**DOI:** 10.1101/2024.12.04.24318451

**Authors:** Tea Pribić, Jayanta K Das, Lovorka Đerek, Daniel W. Belsky, Melissa Orenduff, Kim M Huffman, William E Kraus, Helena Deriš, Jelena Šimunović, Tamara Štambuk, Azra Frkatović Hodžić, Virginia B Kraus, Sai Krupa Das, Susan B. Racette, Nirad Banskota, Luigi Ferruci, Carl Pieper, Nathan E Lewis, Gordan Lauc, Sridevi Krishnan

## Abstract

**Background/Objective:** In a subset of participants from the CALERIE™ Phase 2 study we evaluated the effects of 2y of ∼25% Calorie Restriction (CR) diet on IgG N-glycosylation (GlycAge), plasma and complement C3 N-glycome as markers of aging and inflammaging.

**Methods:** Plasma samples from 26 participants in the CR group who completed the CALERIE2 trial and were deemed adherent to the intervention (∼>10 % CR at 12 mo) were obtained from the NIA AgingResearchBiobank. Glycomic investigations using UPLC or LC-MS analyses were conducted on samples from baseline (BL), mid-intervention (12 mo) and post-intervention (24 mo), and changes resulting from the 2y CR intervention were examined. In addition, anthropometric, clinical, metabolic, DNA methylation (epigenetic) and skeletal muscle transcriptomic data were analyzed to identify aging-related changes that occurred in tandem with the N-glycome changes.

**Results:** Following the 2y CR intervention, IgG galactosylation was higher at 24mo compared to BL (p = 0.051), digalactosylation and GlycAge (the IgG-based surrogate for biological age) were not different between BL and 12mo or BL and 24mo, but increased between 12mo and 24mo (p = 0.016, 0.027 respectively). GlycAge was also positively associated with TNF-α and ICAM-1 (p=0.030, p=0.017 respectively). Plasma highly branched glycans were decreased by the 2y intervention (BL vs 24 mo: p=0.013), but both plasma and IgG bisecting GlcNAcs were increased (BL vs 24mo: p<0.001, p = 0.01 respectively). Furthermore, total complement C3 protein concentrations were reduced (BL vs 24mo: p <0.001), as were Man9 glycoforms (BL vs 24mo: p<0.001), and Man10 (which is glucosylated) C3 glycoforms (BL vs 24mo: p = 0.046).

**Conclusions:** 24-mos of CR was associated with several favorable, anti-aging, anti-inflammatory changes in the glycome: increased galactosylation, reduced branching glycans, and reduced GlycAge. These promising CR effects were accompanied by an increase in bisecting GlcNAc, a known pro-inflammatory biomarker. These intriguing findings linking CR, clinical, and glycomic changes may be anti-aging and inflammatory, and merit additional investigation.

## Introduction

Calorie restriction (CR), a dietary intervention involving a reduction in energy intake below the amount consumed ad libitum, delays aging and increases maximum lifespan in various species^1,2^. Animal studies suggest that CR could extend lifespan and slow aging, however, there are limited data in human populations, and gaps and inconsistencies in our understanding of the mechanisms by which CR is geroprotective^3–6^. Thus, newer mechanisms influencing aging are important to investigate, as they might elucidate human-specific pathways at play^6^.

One potential contributing mechanism is protein N-glycosylation, a post-translational modification. Protein N-glycosylation is crucial to protein folding, stability, and function^7^. Further, with age, protein glycosylation is altered, and these alterations have been linked to various aging-related diseases, such that aberrant N-glycosylation may contribute to aging-related diseases via inflammaging and disruption of proteostasis ^8–10^.

A key example of aging related glycosylation is alterations in glycosylation of IgG. IgG is the predominant antibody (or immunoglobulin) produced by B cells, key players in cellular immunity. With aging, both chronic antigen stimulation and oxidative stress^11,12^ upregulate inflammatory responses leading to chronic low-grade inflammation, or inflammaging, a hallmark of aging^13^. With aging, IgG glycosylation changes include reductions in galactosylation and sialylation^14–19^ and increases in bisecting N-acetylglucosamines (GlcNAc).^20–22^ These aging-related IgG glycosylation alterations are all pro-inflammatory via structural changes that promote an inflammatory response^14–19^. Thus, IgG glycosylation alterations are a result of and accelerate age-related onset of chronic inflammatory disease. Leveraging this, the IgG N-glycome composition and its changes have been used to develop a biological age marker/index^23^. Furthermore, mono and di-galactosylated IgG glycans (but not agalactosylated IgG) and reduced bisecting GlcNAc’s have been negatively associated with incident type 2 diabetes and cardiovascular disease^24–28^.

While not as extensively studied as IgG glycosylation, glycosylation of complement C3 has recently come to be associated with metabolic and autoimmune diseases^29,30^. C3 is a component of the complement cascade, a critical arm of the innate immune system. Greater concentrations of C3 have been noted in metabolic disorders such as adiposity, dyslipidemia, insulin resistance, liver dysfunction, and diabetes, suggesting C3 may be a cardiometabolic risk factor^31,32^. Alternatively, lower C3 plasma concentrations have been tied to longevity in centenarian populations^33^. Aging-related changes in complement C3 include greater amounts of mannose-rich (oligomannose) and oligomannose glucosylated (such as GlcNac2Mannose9Glucose1) glycans. These specific changes are associated with onset of type 1 diabetes and renal complications arising from type 1 diabetes, both of which are hypothesized to be triggered by Endoplasmic Reticulum (ER) stress^30,34^. It is important to note that GlcNAc2 Mannose9Glucose1 is involved in the process of initiating the unfolded glycoprotein response (UPR) which can trigger ER stress^35^. Elevated ER stress signals loss of proteostasis, another hallmark of aging^36,37^ implicating a role for these specific glycoforms in the aging process.

In addition to immune-related glycoproteins like IgG and complement C3, total plasma protein glycome is also linked to age-related disease mechanisms^14^. Longitudinal investigations have identified the relative stability of the total plasma protein N-glycome over several years, while also identifying potential age-related changes^38^. Changes in plasma protein glycosylation patterns, such as appropriate sialylation and galactosylation, mark familial longevity and reflect healthy aging^39^. Moreover, several plasma, serum, and cell surface glycoprotein profiles in human epidemiological, animal model and *in vitro* studies have identified agalactosylation and asialylation signatures, which have now come to be called “glycan” hallmarks of aging^14^.

This manuscript outlines results from a pilot investigation focused on identifying changes in glycomic biomarkers of aging and age-related disease onset in response to a 2y CR intervention in healthy adults with normal weight or mild-moderate overweight (by BMI) from the Comprehensive Assessment of Long-term Effects of Reducing Intake of Energy (CALERIE™) trial. Specifically, IgG, C3, and total plasma protein N-glycosylation were evaluated. We further explored associations between CR-induced changes in a glycome-based biological age clock and epigenetic age clocks. Finally, we investigated muscle transcriptomic changes that are indicative of changes in glycosylation after the 2y CR intervention. Based on our understanding of glycomic changes resulting from aging^17^ and chronic metabolic disease onset^14^ we expected CR to favorably alter IgG and plasma protein N-glycosylation profiles with increased galactosylation and sialylation and reduced branching and bisecting GlcNAc. We expected CR to alter complement C3 glycosylation with less mannosylated and glucosylated C3 glycoforms. Furthermore, we anticipated that these alterations in the N-glycome would correlate with improvements in other biomarkers of aging, such as inflammation and epigenetic age clocks.

## Methods

### Study design

Details of the CALERIE Phase 2 trial were reported previously^40^ and experimental design and protocols archived in on the website of the CALERIE Biorepository (https://calerie.duke.edu/sites/default/files/2022-05/phase2_protocol.pdf). Briefly, this multi-center randomized controlled trial included 220 participants randomized in a 2:1 ratio in favor of CR or to an *ad libitum* (AL) control group^40^. CR group participants were provided with an intensive behavioral intervention and were prescribed an intake 25% below their total daily energy expenditure (which was equal to their energy intake at baseline). Participants’ daily energy expenditure was estimated using doubly labeled water for four weeks at baseline, and subsequent time points to estimate %CR^41^.

### Participants

For this pilot investigation, we selected 26 participants from the CR group who completed the trial and who achieved the highest levels of CR (i.e., ∼10-40% CR) on average.

### Data and biospecimens from biorepository

Plasma samples from the CALERIE2 study were obtained after approval from the CALERIE Steering Committee and NIA AgingResearchBiobank. De-identified plasma samples were recoded to blind the study personnel conducting analyses to the study timepoints. These samples are available from the NIA AgingResearchBiobank upon request (https://agingresearchbiobank.nia.nih.gov/studies/calerie/) and were obtained for baseline, 12 mo and 24 mo. In this analysis we included anthropometrics (age, BMI, body weight, body composition), energy intake, energy expenditure and %CR, immune biomarkers (plasma TNF-α IL6, IL8, MCP1 (Monocyte chemoattractant protein-1), PDGFAB (platelet-derived growth factor AB), TGFB1 (transforming growth factor – β1), CRP) and clinical variables (plasma HDL, LDL, triglycerides, total cholesterol, cortisol, fasting glucose and insulin, Homeostatic Model Assessment of Insulin resistance, HOMA-IR, Homeostasis Model Assessment of β-cell function (HOMA-β), Insulin-like growth factor binding protein-1 and -3 (IGFBP1 and IGFBP3)) that were collected as part of the study. In addition, DNA methylation-based epigenetic clock information previously published^42^ (available for all n = 26 participants) was analyzed to evaluate associations with glycomic biological age measures. Finally, skeletal muscle transcriptome data published earlier^43^ measured for n = 75 participants were also analyzed to identify glycosylation gene changes that were observed with the CR intervention.

### Total plasma protein N-glycome analysis

For N-glycan release from total plasma proteins, each plasma sample (10 μL) was aliquoted into 1 mL 96-well collection plates (Waters, Milford, MA, USA) followed by a deglycosylation (release of glycans) procedure. Plasma proteins were denatured by the addition of 20 μl 2% (w/v) sodium dodecyl sulfate (SDS, Invitrogen, Carlsbad, CA, USA), and the sample was incubated at 65°C for 10 minutes. After denaturation, 10 μl of 4% (v/v) Igepal-CA630 (Sigma Aldrich, USA) was added to the samples, and the mixture was shaken for 15 minutes on a plate shaker (GFL, Germany). N-glycans were released by adding 1.2 U PNGase F (Promega, USA) and incubating overnight at 37 °C.

### Fluorescent labeling and HILIC-SPE clean-up of released *N*-glycans

Released plasma N-glycans were labelled with 2-aminobenzamide (2- AB, Sigma-Aldrich). The labeling mixture consisted of 0.48 mg 2 AB and 1.12 mg 2-picoline borane (2-PB, Sigma-Aldrich) in 25 μl dimethyl sulfoxide (DMSO, Sigma-Aldrich) and glacial acetic acid (Merck, Darmstadt, Germany) (7:3, v/v) per sample. The labeling mixture was added to each sample followed by incubation at 65 °C for 2 hours. Excess reagents and proteins were removed from the samples using solid phase extraction by hydrophilic interaction liquid chromatography (HILIC-SPE) using 0.2 μm wwPTFE 96-well membrane filter plates (Pall, New York, NY, USA). The N-glycans were eluted with water and stored at - 20 °C until use.

### Hydrophilic interaction liquid chromatography of *N*-glycans

2AB-Fluorescently labelled glycans were separated on a Waters Glycan Premier BEH amide chromatography column, 150 × 2.1 mm x 1.7 μm BEH particles (Waters, USA), with 100 mM ammonium formate, pH 4.4 as solvent A and ACN as solvent B. A linear gradient of 30-47% ACN (Honeywell, USA) (v/v) at a flow rate of 0.561 ml/min was used for separation in a 32.5 min analytical run. The excitation and emission wavelengths were set to 250 nm and 428 nm, respectively. The chromatograms obtained were separated into 39 peaks (GP1-GP39) (Supplemental Figure 1). The glycan peaks were analyzed by their elution positions and measured in glucose units, which were then compared to the reference values in the "GlycoStore" database (available at: https://glycostore.org/) for structure assignment ^44,45^. The amount of glycans in each peak was expressed as a percentage of the total integrated area. For plasma glycans, 16 derived features were calculated in addition to 39 directly measured glycan features. (**Supplemental Table 1**).

### Immunoglobulin G (IgG) N-glycome analysis

#### Isolation of IgG from human plasma

Samples were randomly distributed across three 96-well plates. To isolate IgG from a 25 μL plasma sample, the CIM^®^ r-Protein G LLD 0.05 mL Monolithic 96-well plate (2 µm channels) (BIA Separations, Slovenia, Cat No. 120.1012-2) was used according to published protocols^17^. The vacuum-assisted setup utilized a manual system consisting of a multichannel pipette, a vacuum manifold (Pall Corporation, USA), and a vacuum pump (Pall Corporation, USA), with pressure- reductions of approximately 5 mmHg during sample application and IgG elution. Following the isolation of IgG, the monolithic plate was stored in 20 % (v/v) EtOH in 20 mM TRIS + 0.1 M NaCl, pH 7.4 at 4 °C. Subsequently, 20 μL of IgG was aliquoted in a PCR plate (Thermo Scientific, UK) and dried in a vacuum centrifuge. For IgG N-glycan analysis by capillary gel electrophoresis with laser-induced fluorescence (CGE-LIF), deglycosylation, released N-glycan labeling and clean-up of labeled IgG N-glycans were performed using modified protocols described elsewhere^17,46^.

#### IgG N-glycan release, labeling, and clean up

Isolated IgG samples were dried in a Savant SpeedVac vacuum concentrator, resuspended in 3 μL of 1.66 x PBS (w/v), and denatured in 4 μL 0.5% SDS (w/v) (Sigma-Aldrich, USA), followed by incubation at 65 °C for 10 minutes. Following incubation, 2 μL of 4% Igepal-CA630 (Sigma-Aldrich, USA) was added to the samples and incubated on a shaker for approximately 5 minutes. For glycan release, 1 μL of the enzyme mixture (1.2U of PNGase F (Promega, USA) in 1 μL 5× PBS) per sample was added and incubated for 3 hours at 37 °C. After incubation, the deglycosylated IgG mix was dried in a vacuum concentrator for 1h, and diluted with 2μL of ultra-pure water, and left on a shaker for approximately 5min to prepare the labeling mixture for labeling the released N-glycans. The labeling mixture was freshly prepared by combining 2 μL of 30 mM 8-aminopyrene-1,3,6-trisulfonic acid trisodium salt (APTS) (Synchem, DE) in 3.6 M citric acid (Sigma-Aldrich, USA) with 2μL of 1,.2 M 2-picoline borane in dimethylsulfoxide (Sigma-Aldrich, USA) per sample. Mixing was achieved by vortexing several times, followed by a 16h incubation at 37 °C. The labeling reaction was stopped by adding 100 μL of cold 80% acetonitrile (Carlo Erba, Spain).

For the clean-up procedure, 200 μL of Bio-Gel P-10 slurry per well was added on a 0.2 μm wwPTFE AcroPrep filter plate (Pall Corporation, USA) and used as the stationary phase. The wells containing Biogel P-10 were prewashed with 200 μL ultrapure water and 200 μL 80% cold ACN (v/v) (Carlo Erba, Spain), three times for each step. The samples were loaded into the wells and after a 5-minute incubation on a shaker, samples were subsequently washed 5 × with 5 × 200 μl 80% ACN containing 100 mM TEA, pH 8,5 (Carlo Erba, Spain/Millipore Sigma, USA) followed by 3 x 200 μl 80% ACN (Carlo Erba, Spain) after 2-minute of incubation at room temperature. IgG N-glycans were eluted with a total of 500μl of ultrapure water after 5-minute incubation at room temperature after each step, and combined eluate fractions were stored at –20°C until use.

### Capillary gel electrophoresis N-glycan analysis and data processing

The IgG N-glycoprofiling by capillary gel electrophoresis with laser-induced fluorescence (CGE-LIF), was performed using an Applied Biosystems 3500 Genetic Analyser (Thermo-Fischer Scientific, USA)), equipped with 50 cm long 8-capillary array (Thermo-Fischer Scientific, USA). Polymer POP-7 (Thermo-Fischer Scientific, USA) was used as a separation matrix in capillaries.

In a 96-well MicroAmp Optical 96-Well reaction plate, 2μl of APTS-labeled N-glycans were combined with 8μl of HiDi formamide, ensuring a thorough and uniform resuspension. The instrumental method was created by setting the operating parameters as follows: injection time: 9s; injection voltage: 15kV; run voltage: 19.5kV; oven temperature: 60°C; and working time: 1000s. The obtained electropherograms were integrated in the same manner into 27 peaks using Waters Empower 3 software. The Glycan composition of each peak has been previously determined^17^. For IgG glycans, 12 derived features were calculated in addition to 27 directly measured glycan peaks (**Supplemental Table 2**).

### IgG-glycan based biological age calculation

An IgG-glycan based biological age index was developed previously by analyzing plasma glycome from ∼5100 individuals from diverse populations using a linear gradient separation method^23^. In total 906 individuals from the Croatian island Vis (377 men, 529 women), 915 individuals from the Croatian island Korcula (320 men, 595 women), 2,035 individuals from the Orkney Islands in Scotland (797 men, 1238 women), and 1,261 female twins from the TwinsUK cohort were measured. Their age ranges varied across cohorts, 16 to 100 years in Orkney, 18 to 98 in Korcula, 18 to 93 in Vis, and 27 to 83 in the TwinsUK cohort. The biological age index was developed as a function of specific galactosylated glycans (GP6, GP14 and GP15 – sialylated, mono and di-galactosylated low-branched glycans) and were found to explain a greater proportion of variance in biological age than telomere length measures. In particular, the lesser galactosylated IgG’s were identified to be a strong predictor of biological and chronological age, and inflammaging, while the more galactosylated versions reflected the opposite. These findings establish the IgG glycome as a marker of both inflammaging, and aging-related diseases. Since its development, either the IgG-glycan based biological age index or its chromatogram components (GPs) have been evaluated in various conditions including diabetes^47^, cardiovascular disease^48^, rheumatoid arthritis^49^, and neurodegenerative diseases^50^ and demonstrated strong disease predictive capability. Using this^23,51^, an IgG-glycan-based biological age (GlycAge) was calculated with one deviation from the reported approach. The current analysis included a final range-scaling step (as shown below) to ensure that GlycAge was within the range of the participants in the CALERIE2 study.

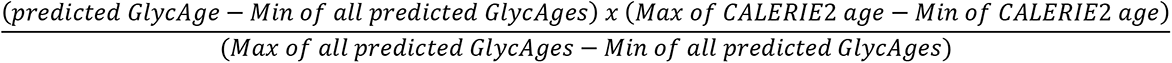

### Complement component (C3) N-glycome analysis

#### Lectin-based C3 enrichment

Complement component C3 was enriched from human plasma following the published method^34^ with minor adjustments. Briefly, Concanavalin A (Con A)-Sepharose 4B resin (7.5 µL) was preconditioned with a binding buffer (20 mM Tris-HCl, 0.5 M NaCl, pH 7.4) on 96-well polypropylene filter plate (Orochem Technologies Inc). Plasma samples (10 µL) diluted with the binding buffer in ratio 1:9 were loaded on conditioned resin and incubated for 2 hours at 4 °C, while shaking. Matrix was washed with 250 µL of binding buffer three times and incubated in 200 µL of elution buffer for 45 min before glycoproteins were eluted and dried in the vacuum concentrator.

#### C3 digestion and glycopeptide enrichment

Dried samples were resuspended in 78 µL of 15% (v/v) 2-propanol in 0.1 M ammonium acetate and denaturated at 60 °C for 10 min. Subsequently, 2 µL of endoproteinase Glu-C (0.5 U/ µL) was added to samples for the incubation overnight at 37 °C. Hydrophilic interaction chromatography based solid phase extraction (HILIC-SPE) enrichment of glycopeptides was done as previously described^34^ using 85 % acetonitrile (ACN) instead of 90 %.

#### Nano-LC-ESI-MS analysis

Enriched C3 glycopeptides were analysed using an Ultimate 3000 RSLC nano system (Dionex/Thermo Fisher Scientific) coupled to Orbitrap Exploris 240 (Thermo Fisher Scientific) using an EASY-Spray source. Samples (1 µL) were loaded onto Acclaim PepMap C18 trap column (5 × 0.3 mm, 5 μm, 100 Å, Thermo Fisher Scientific) at a flow 80 µL/min and washed for 3 min. Separation was performed on EASY-Spray PepMap™ RSLC C18 column (75µm x 150 mm, particle size 3 µm, pore size 100 Å, Thermo Scientific) at the flow of 0.5 µL/min with a linear gradient. The column was maintained at 35 °C. The MS instrument was operated in a data-dependent mode. Samples were ionized at a spray voltage of 2250 V and full MS1 scans were acquired within *m/z* 600-2,000 with a 60,000 resolution and standard AGC target. The RF level was set to 90% and maximum injection time to automatic. Previously annotated glycopeptides separated based on the peptide backbone in two clusters with three glycoforms each. Data were extracted and quantified using LacyTools software^52^.

#### Immunoturbidimetric C3 assay

The plasma concentration of complement C3 was measured using an immunoturbidimetric method with a commercial reagent kit (Beckman Coulter Inc., Brea, CA, USA) on the AU5800 Series Clinical Chemistry Analyzer (Beckman Coulter Inc., Tokyo, Japan) according to the Manufacturer’s instructions. C3 from the sample reacts with the specific antibody from the reagent and forms insoluble complexes. The quantity of formed complexes is monitored spectrophotometrically and the absorbance of the complex is proportional to the C3 concentration in the sample. The reference range was 0.79–1.52 g/L.

## Statistical analysis

Data analyses were done using JMP Pro 17.2.0 (SAS Institute, Cary NC) and R Statistical Software (R Studio version 2023.06.1) ^53^. All data were evaluated for normality and missingness. Data were transformed to log, cube root or Johnson family of distributions to achieve normality prior to analysis, evaluated using Q-Q plots and Shapiro-Wilk tests. For anthropometric parameters, repeated measures mixed model analysis of covariance was used to evaluate the differences between baseline (BL), mid-intervention (12 mo) and end of intervention (24 mo) using participant as random effect. For all glycan parameters, repeated measures mixed model analysis of co-variance was used to evaluate the differences in glycan parameters between baseline, 12 mo and 24 mo. using participant as a random effect, baseline, age and sex as covariates, followed by FDR adjustment for multiple comparison correction. Where appropriate, family-wise multiple comparison tests (Tukey’s) were performed to obtain individual time point differences. Bivariate regression with repeated measures, and repeated measures correlation (package rmcorr in R^54^) were used to evaluate relationships between glycan parameters and other features (epigenetic clocks from Waziry et al^42^, and immune and metabolic parameters) to account for repeated measures from the same individual at multiple time points. There was missing data in immune and metabolic parameters at baseline, and analyses in these instances were done using only available timepoints. Skeletal muscle differential gene expression (DGE) analysis was reported by Das et al ^43^ comparing 57 CR and 33 AL group individuals from the CALERIE2 trial. The DGE identified the top 1003 genes whose expression changed differentially between BL and 24mo at p<0.05. Here, we further identified mammalian glycosylation genes (glycogenes) encoding glycosylation machinery enzymes as listed in Glycoenzyme repository (https://glycoenzymes.ccrc.uga.edu/) that were in the 1003 identified genes from the DGE. Functional enrichment analysis using only the glycogenes was done with g:Profiler, which identified specific glycosylation processes that were different between the CR and AL group.

## Results

In the 26 participants included in this pilot study, % CR was ∼18% in the BL–12mo interval, and went down to ∼9% in the 12-24mo interval (Fig 1). Body weight reduced significantly between BL and 12mo, and increased slightly from 12-24mo.

**Figure 1.**
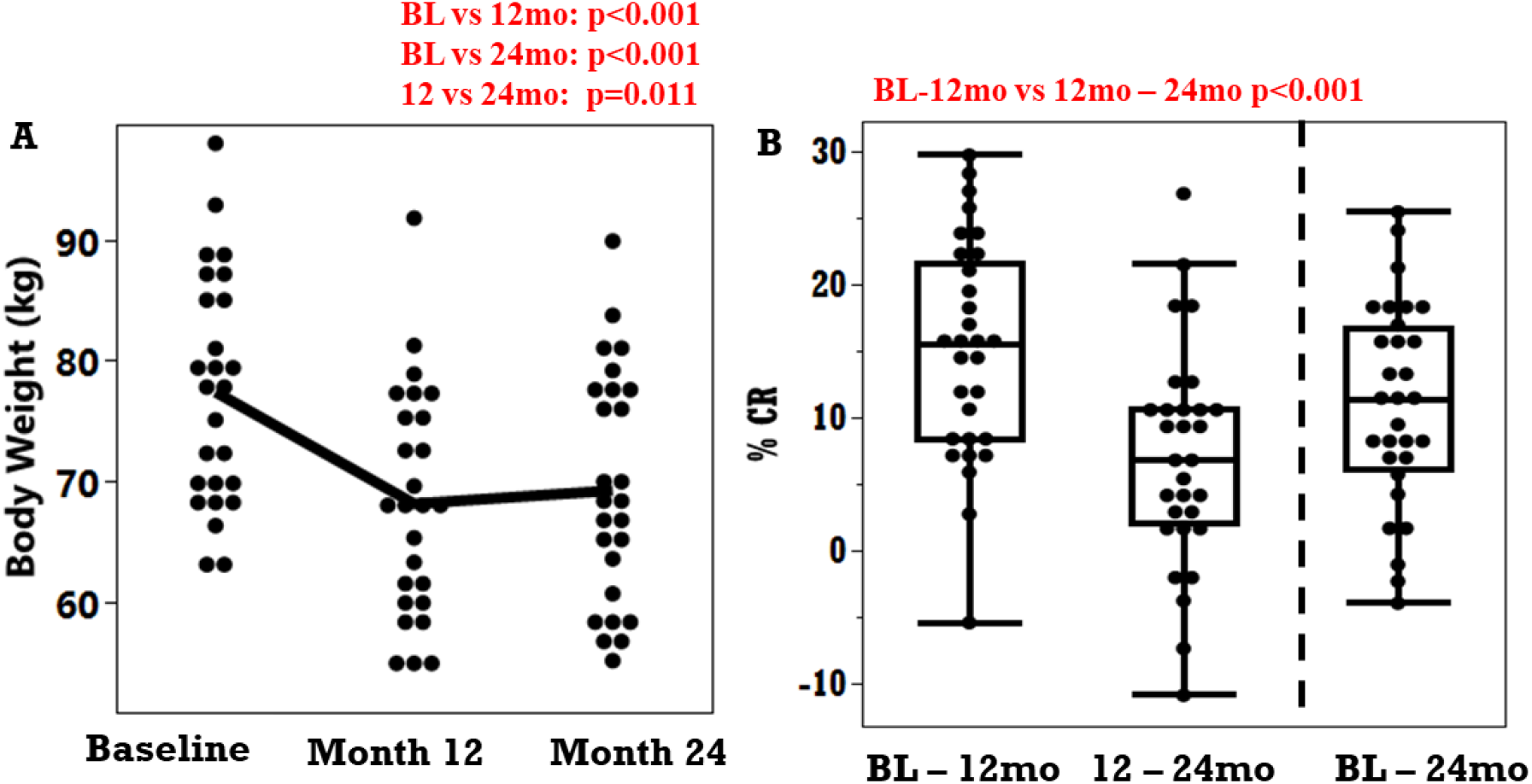
Anthropometric and calorie restriction characteristics of study subset of participants (n = 26, 13 male, 13 female) included in this report. **A:** The box and whisker plots depict body weight and **C:** energy intake at baseline (BL), 12 month and 24 month. In **B**: the dotted line separates the individual 12mo intervals from the total study %CR, and the plots depict %CR during each intervention period. Inset p values for A and C were obtained from mixed models using months as repeated measure and participant as random effect, for B were obtained from mixed models that used time intervals as repeated measures and participants as random effects.

## CR effects on GlycAge Index of biological age

A comparison of the chronological age of participants in this cohort (25.6 - 49.9y at baseline) and their calculated GlycAge is presented in **Figure 2**. At 12mo, GlycAge was not different from chronological age, but at 24mo, GlycAge was lower than chronological age (Figure 2A). A comparison of the slopes of a linear regression fit between GlycAge and chronological age against months in study (BL-12mo and 12-24mo) identified that the slope of GlycAge change between 12-24mo was significantly different from that of chronological age, but not between BL-12mo. Mean GlycAge (Figure 2B) decreased between 12mo and 24mo (p = 0.027) but not between BL and 12mo or between baseline and 24 mo. However, there was large inter-individual variability that warrants further investigation. During the BL–12mo interval (when the GlycAge change tracked chronological age), change in body weight was inversely associated with change in GlycAge; however, in the 12–24mo interval (when GlycAge was reduced in response to the intervention, there was no association between body weight change and GlycAge change. A summary of all outcomes presented in this report are tabulated in Supplemental Table 3.

**Figure 2.**
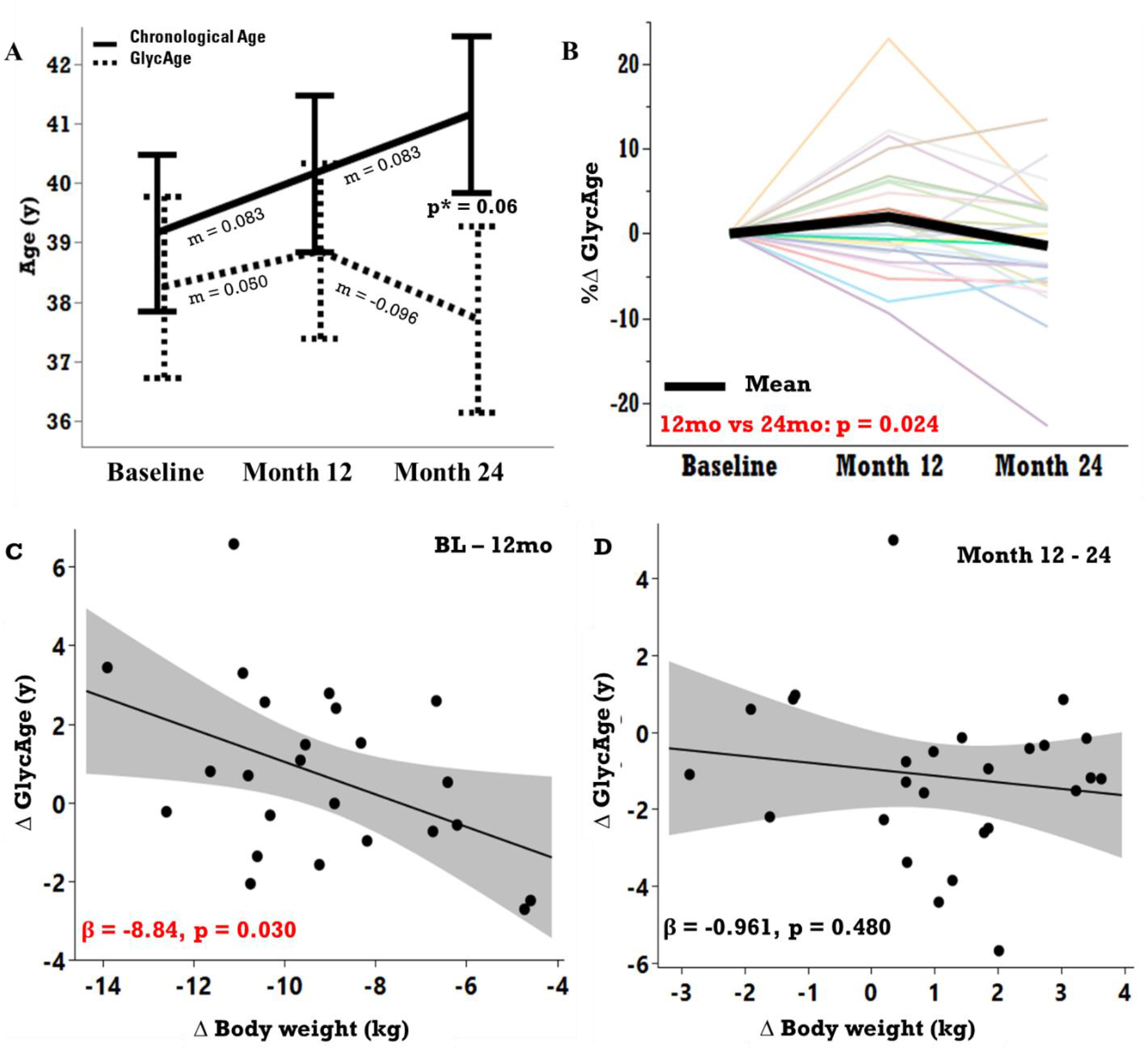
IgG based biological age GlycAge changes significantly from the intervention. **A.** Chronological age compared to IgG based biological age (GlycAge), with the slope (m) of each time interval (0 – 12mo and 12-24mo) inset, which were different (p=0.009). However, a multiple comparison test (Tukey’s) comparing the two ages indicated that at 24mo the two ages showed a difference of moderate significance (p=0.06). **B.** Biological age (IgG based GlycAge) percent change from baseline with significant p-value inset. Differences were evaluated using a mixed model using visit month as repeated measure, participant as random effect, sex and baseline GlycAge as covariates followed by FDR correction. If the FDR corrected p-values were significant, multiple comparison tests (Tukey’s) were conducted. Tukey’s tests indicated that the 12mo – 24mo ages were different (p=0.027). **C and D**. Change from baseline visit at 12mo and 24mo of IgG based biological age GlycAge. Bivariate regression plots are shown with the β-estimate and corresponding p-value inset evaluating the relationship between change in body weight and GlycAge by time interval (BL-12mo and 12-24mo).

## CR effects on the IgG N-glycome

Compared to 12mo, IgG di-galactosylation was greater at 24mo (p = 0.039). Compared to BL, total galactosylation (which is the sum of glycan structures with single and double galactoses- Fig 3C) was greater at 24mo (p = 0.051). While there was no significant change in agalactosylation (i.e. glycan structures with no galactose sugars, Fig 3D, *p = 0.073*), bisecting GlcNAc was higher at both 12mo and 24mo compared to BL (p <0.001 and 0.019 respectively, Fig 3E).

**Figure 3.**
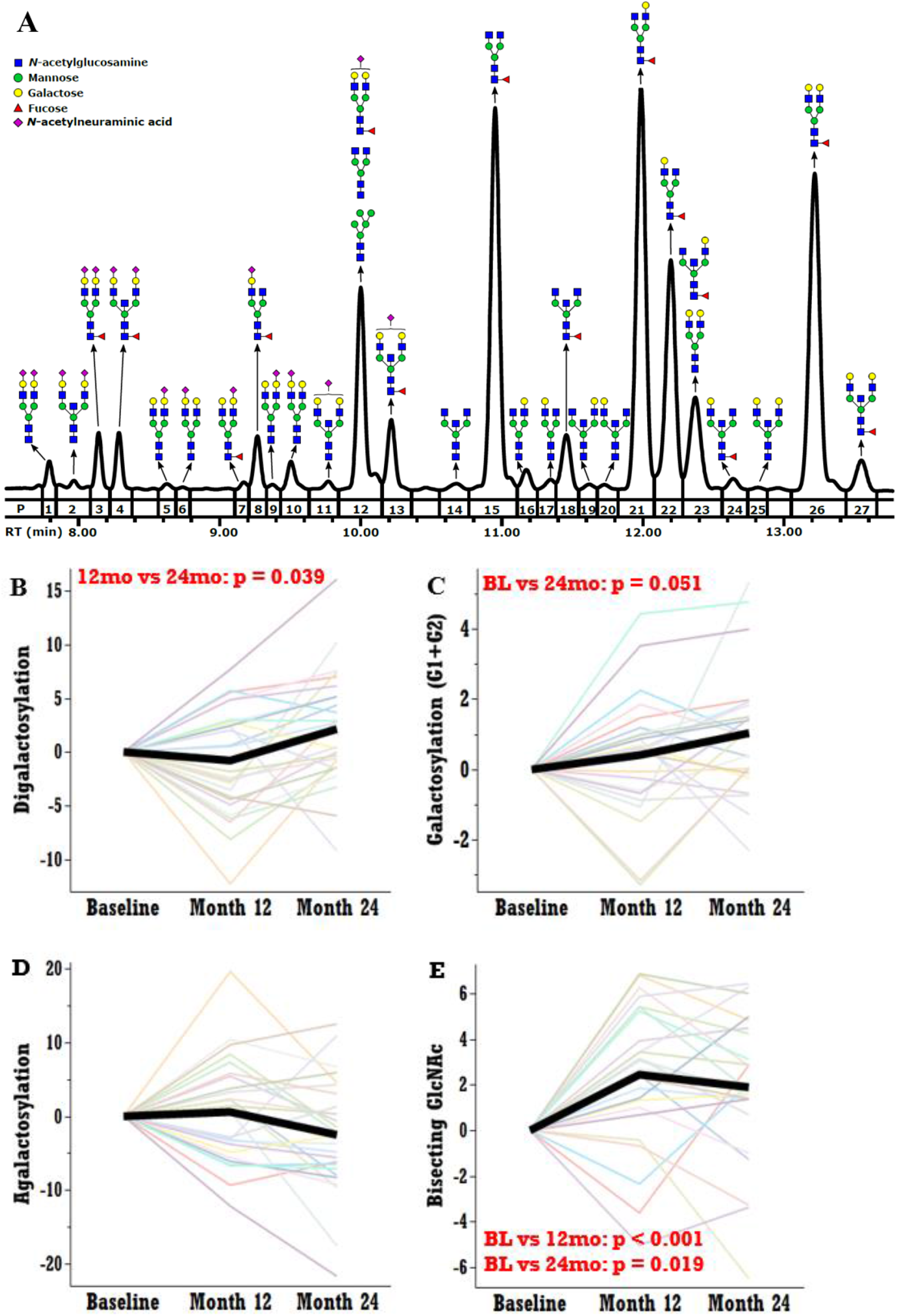
IgG glycome summary from CALERIE. **A.** CGE-LIF electropherogram of IgG glycome showing different glycans detected along with the key indicating their structures. **B, C, D and E** show percent change from baseline of IgG glycome in a subset of CALERIE study participants (n = 26). Data were evaluated by mixed models using visit month as repeated measure, participant as random effect, baseline values for each parameter, sex and age as covariates followed by FDR correction. If the FDR corrected p-values were significant, multiple comparison tests (Tukey’s) were conducted. Significant p values (inset in red) were obtained from Tukey’s tests.

## CR effects on total plasma N-glycome

Changes in select plasma glycome features before and after the 2y CR are presented in **Figure 4**. Of particular interest are (a) sialylated and galactosylated glycans, both typically reduced with aging and chronic inflammation, (b) highly branched glycans, usually found at greater concentrations in chronic inflammatory diseases, and (c) bisecting GlcNAcs (a sub-type of branched glycan), which are also known to increase with biological age and chronic inflammation^14^. At 24 mo, concentrations of tetra-sialylated glycans were reduced compared to BL (p < 0.001) and 12mo (p = 0.031). On the other hand, asialylated glycans were higher at 24mo compared to BL (p=0.011) and 12mo (p= 0.036).

**Figure 4:**
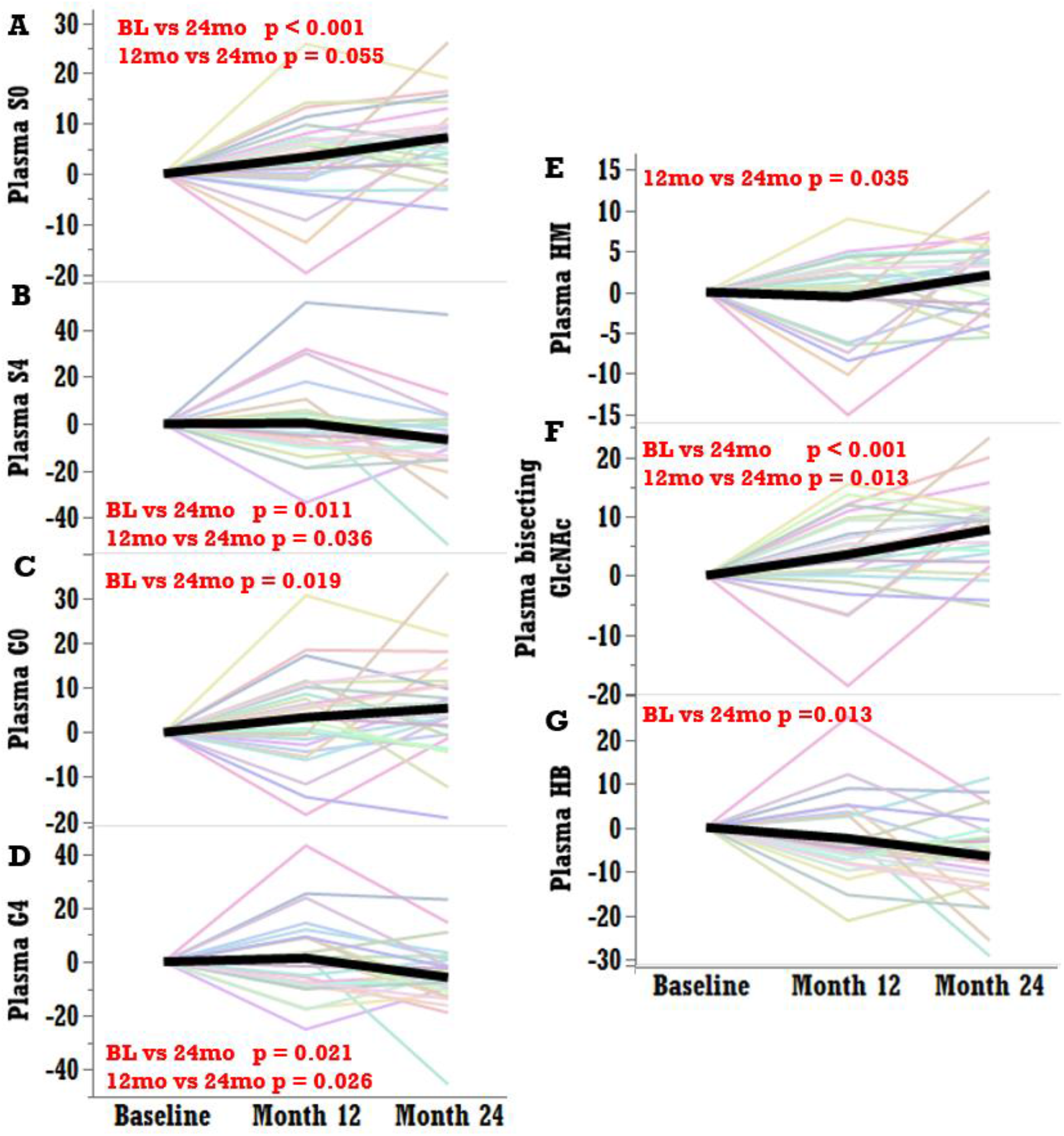
Plasma glycome effect of 24 mo. CR evaluated by mixed model tests using visit month as repeated measure, participant as random effect, baseline parameters, sex and age as covariates followed by FDR correction. If the FDR corrected p-values were significant, multiple comparison tests (Tukey’s) were conducted and significant p values, if any were inset in red. All figures show percent change from baseline. A and B Non-sialylated glycans were higher and tetra-sialylated glycan species were lower at 24mo compared to BL. C and D Non-galactosylated species did not significantly change, however, tetra- galactosylated glycans reduced in the 12-24mo period of CR. E oligomannose glycans were significantly different in the mixed model, but Tukey’s multiple comparison tests found BL vs 24mo and 12 vs 24mo only moderately different (p∼0.08 for both). F Bisecting GlcNAc in plasma were higher at 12mo and 24mo compared to BL. G Highly branched glycans were, however, lower at 24mo compared to BL.

For the total glycome, from BL to 24mo, there was an increase in agalactosylated glycans (p = 0.019), a reduction in tetra-galactosylated (p = 0.021) and highly branched glycans (p = 0.013). There was also an increase from BL to 12- and 24mo in bisecting GlcNAcs (p < 0.001, and p = 0.013, respectively). Oligomannose glycans (HM) were also higher at 24mo compared to 12 mo (p = 0.035).

## CR effects on Complement C3 N-glycome

Compared to baseline, at 12 and 24mo, Complement C3 protein concentrations were significantly lower (p < 0.001, p < 0.001 respectively, **Figure 5**). Among the C3-Asn85 glycoforms (glycosylated at the 85^th^ Asparagine residue), compared to baseline, the Man5 (Asn85 N2H5) glycoform was reduced at 24mo (p = 0.002) and the Man6 (Asn85 N2H6) glycoform was elevated at 24mo (p<0.001). The Man7 (Asn85 N2H7) glycoform was lower at 12mo compared to baseline (p<0.001) and higher at 24mo compared to 12mo (p < 0.001).

**Figure 5:**
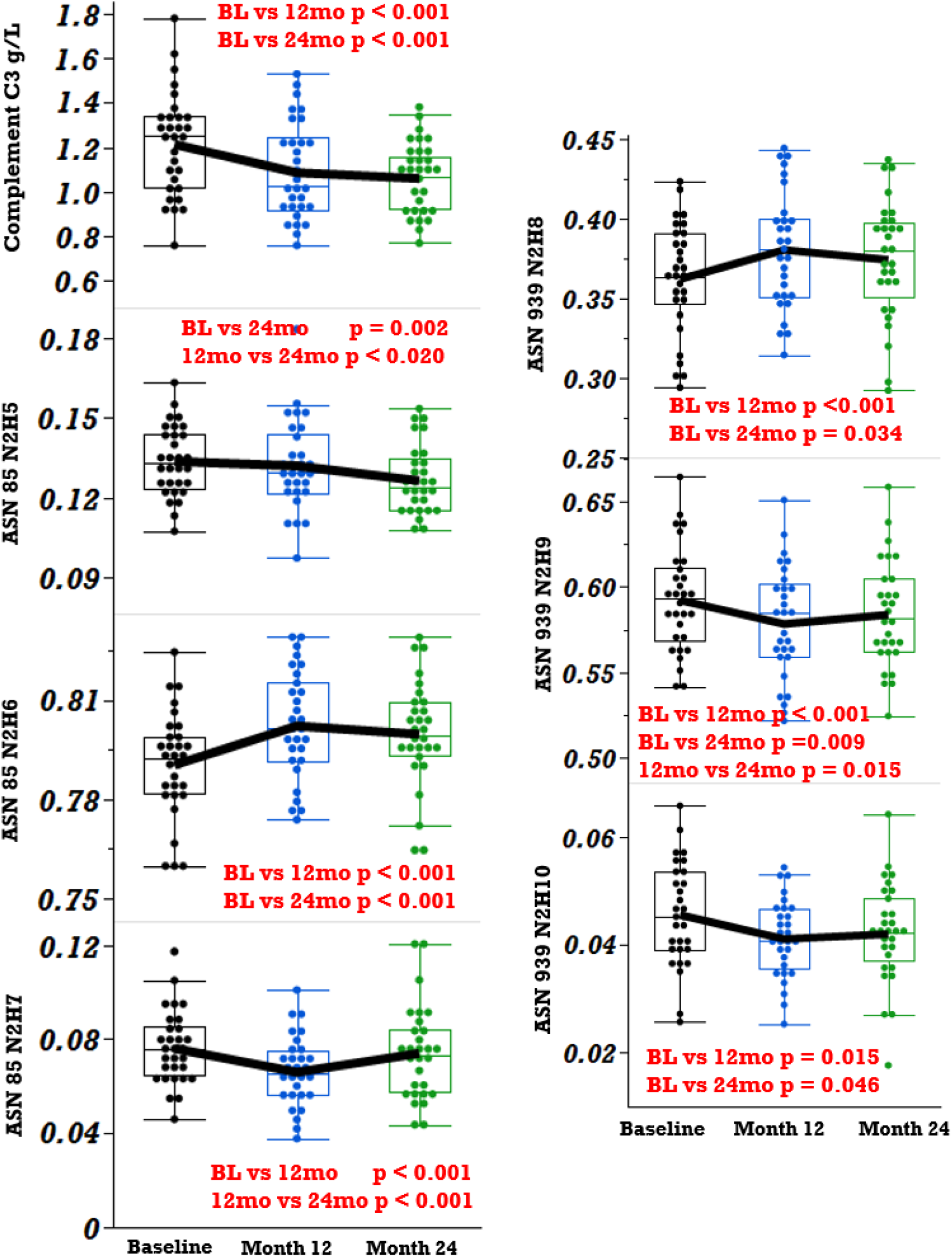
Complement C3 and C3 glycans before and after the 2y CR protocol in CALERIE2. All box plots show concentrations of total protein C3 and C3glycans at baseline, 12mo and 24mo of the CALERIE study in n = 26 participants. Data were evaluated using mixed models using visit month as repeated measure, participant as random effect, baseline values, sex and age as covariates followed by FDR correction. If the FDR corrected p-values were significant, multiple comparison tests (Tukey’s) were conducted and significant p values, if any were inset in red.

Among the C3-Asn-939 glycoforms (glycosylated at the 939th Asparagine residue), compared to baseline, at 12 and 24 mo, the Man8 glycoform (Asn939 N2H8) was greater (p <0.001, p = 0.034 respectively), while Man9 (Asn-939 N2H9) was reduced at 12 (p<0.001) and 24mo (p = 0.009). Compared to baseline, Man9Glc1 (Asn939 N2H10: Glucose1Mannose9GlcNAc2) glycoforms were decreased at 12 and 24mo (p = 0.015, p = 0.046 respectively).

## Association between GlycAge and other epigenetic clocks

Using this subset of n=26, Figure 6A depicts a comparison of chronological age to previously published epigenetic clock data from CALERIE^42^ as well as the GlycAge index. Upon visual inspection, like the complete study cohort, in this smaller subset, DunedinPACE appears to be reduced by CR while other epigenetic clocks appear to indicate increased age with CR. No other positive (or inverse) significant correlations were observed between GlycAge and other epigenetic clocks or DunedinPACE (Fig 6G).

**Figure 6:**
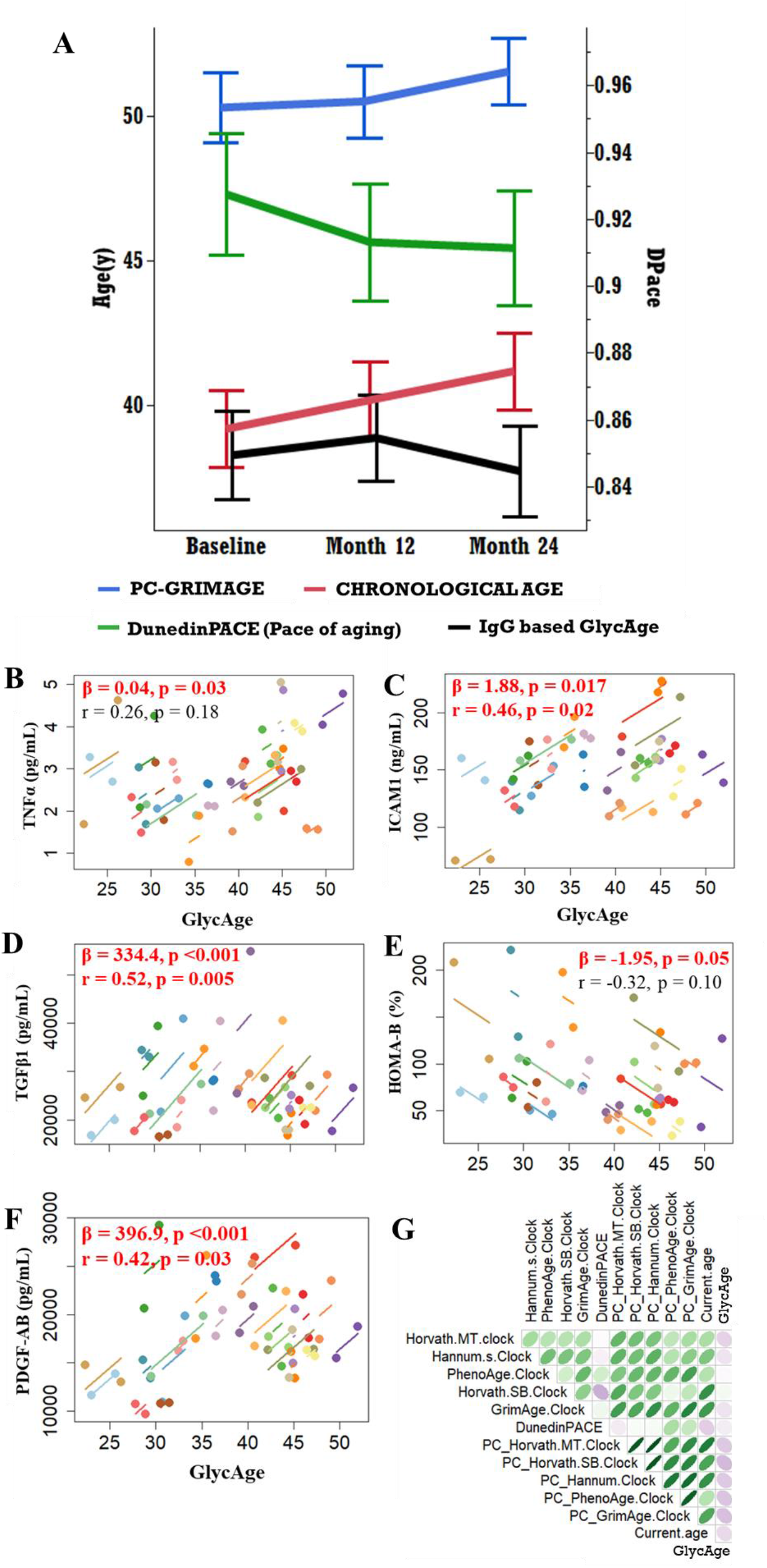
Relationship between GlycAge, other biological age clocks and immune parameters. A. Chronological age, GlycAge, PC-GrimAge and DunedinPACE at Baseline, 12 and 24mo of the intervention. B, C, D, E and F. Repeated measures correlations are shown (each color is one participant from n = 26) between GlycAge and select immune parameters of interest at baseline, 12 and 24 mo. Correlation coefficient r and corresponding p values are shown, along with a repeated measures bivariate regression β-estimate and corresponding p-value. G. A correlation matrix also depicting repeated measures correlation coefficients between several epigenetic ages, GlycAge and chronological age at months 12 and 24 (these measures were not collected at month 0). PC – principal components; SB - skin and bone, MT - multi-tissue

## Associations between GlycAge and immune/metabolic parameters

Repeated measures correlation analysis revealed that GlycAge was significantly positively correlated with TGF-β, ICAM1, PDGF-AB and TNF-α, while being inversely associated with HOMA-B (Figure 6B, C, D, E and F).

## Skeletal muscle transcriptomic changes in glycosylation processes due to CR

Figure 7 shows transcriptional changes in skeletal muscle glycogenes; CALERIE2 global DGE data were previously reported^43^. Compared to BL, at 24 mo, muscle expression was greater for CHST3, a heparan sulfate processing enzyme (p<0.01) and HSPA5 an ER resident Hsp70 chaperone^55^ involved in UPR (p<0.05) (Fig 7A). The largest number of differentially expressed genes in glycogene-related processes occurred in the ER and Golgi body, as expected, since the ER and the Golgi apparatus are responsible for most glycosylation processes (Fig 7B). While N- and O-glycosylation processes were only ∼1% of the total changes observed, glycosaminoglycans (GAGs) were ∼9%.

**Figure 7:**
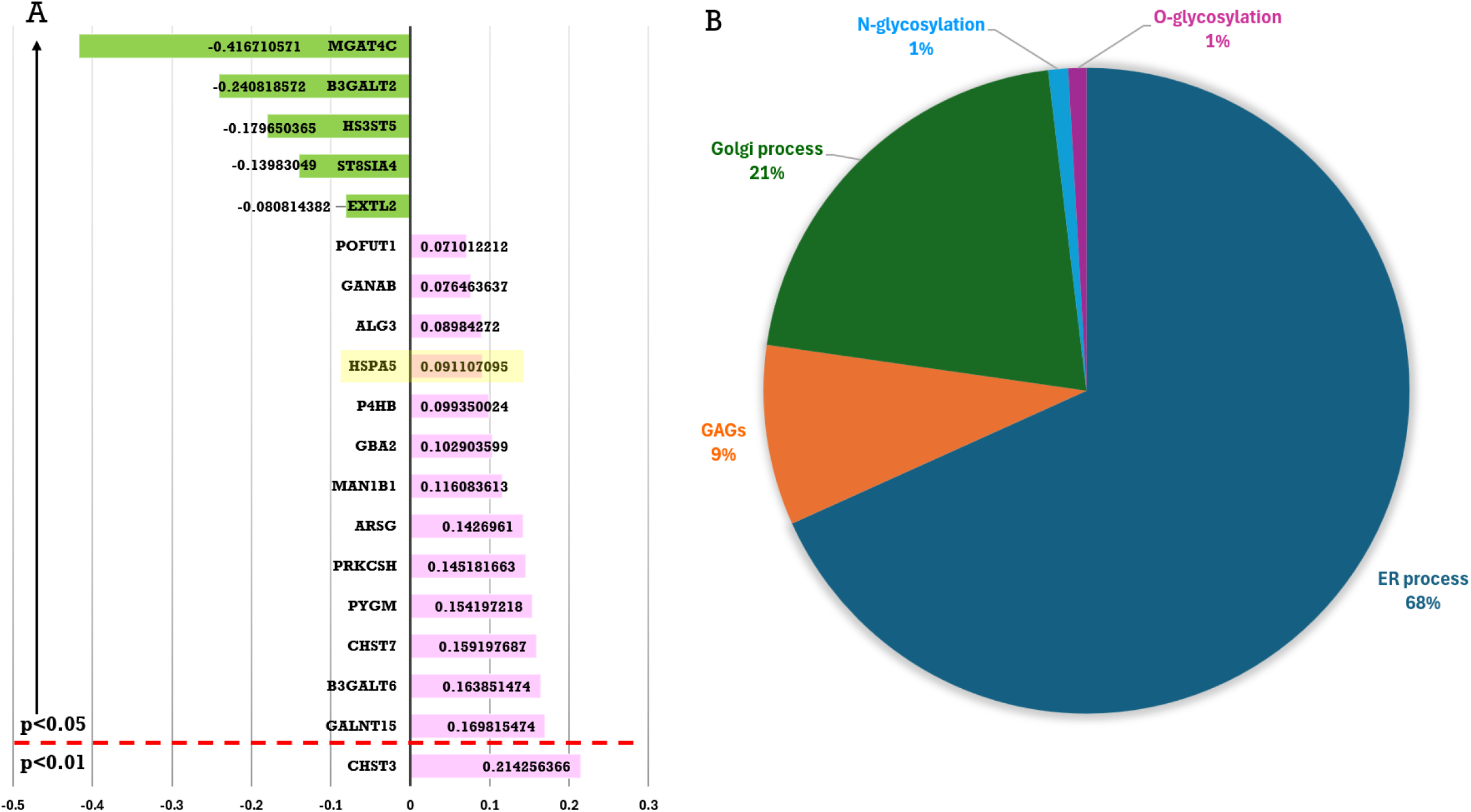
Differential gene expression analysis (DGE) of muscle (vastus lateralis) transcriptome data in n = 90 (57 CR and 33 controls). A shows glycogenes that were differentially expressed over the two-year CR (linear) at p<0.05 threshold (CHST3 is the exception with a p value of <0.01). The standardized beta values plotted, and listed, are positive if their expression was higher in CR at 24mo compared to baseline and inverse if lesser at 24mo compared to baseline. B depicts the percent of differentially-expressed secretory pathway-related genes that are associated with different glycosylation processes, that are overexpressed in CR based on beta values in A. GAGs – glycosaminoglycans/proteoglycans, ER – endoplasmic reticulum.

## Discussion

In this pilot study of 26 participants who underwent 24 months of CR, detailed glycosylation profiling indicates CR increased IgG galactosylation, reduced highly branched and highly sialylated glycans, reduced Complement C3 protein and highly mannosylated and mannosylated and glucosylated glycans on C3, along with a reduction in GlycAge. Furthermore, GlycAge changes were associated with a few other established inflammation- and aging-related biomarkers, demonstrating the potential of glycans in assessing the impact of CR on biological ageing. In addition, skeletal muscle transcriptional analyses suggest glycosylation changes result from CR impacts on the ER at the transcriptional level. Specifically, after 2y of CR, skeletal muscle expression of HSPA5, the binding immunoglobulin protein coding gene (involved in UPR), was upregulated.

With CR, GlycAge, an IgG glycan-based clock of biological age, was reduced from 12 mo to 24 mo, during the period of weight stability. GlycAge was not associated with biological age based on any of the other epigenetic clocks. However, in this subset of 26 participants, none of the examined epigenetic clocks were reduced by CR intervention, except for DunedinPACE, which approximates a pace of aging^42^. Consistent with an inflammaging biomarker^56^, GlycAge was positively associated with various inflammatory biomarkers (TGFβ1, TNFα, PDGF-AB and ICAM1), which are known to increase with age and aging-related diseases^57^. CR also led to increase in IgG galactosylation, an IgG glycan feature usually associated with younger age and IgG’s anti-inflammatory potential^58^. Surprisingly, an increase in IgG bisecting GlcNAc was also observed, a feature usually associated with inflammation^21^, suggesting that while some aging related changes in the glycome are affected by CR, others may not be.

Our findings are consistent with others, where extensive weight loss induced by a 3-week low-calorie diet, followed by bariatric surgery, has been shown to alter IgG glycosylation and reduce inflammageing^59^. In the DIOGENES study^60^ (n=1850) the 8-week weight loss period reduced IgG agalactosylation, but did not affect galactosylation. However, at the end of the 6- month weight maintenance period, there was an increase in IgG digalactosylation, and an increase in bisecting GlcNAc. Similarly, in the CALERIE2 study, the initial baseline – 12 mo period, when the weight loss was maximal (∼-10kg), IgG digalactosylation and GlycAge did not change significantly, but did change during the 12 mo to 24 mo period, when weight loss was minimal (∼+0.5). While the %CR in the 12-24mo period recorded was lower, there was still substantial CR observed in the study population^61^ and in this cohort, as observed in Fig 1B that shows the average CR during the entire 24mo duration. As noted earlier, % CR decreased within the first 20weeks (5 months) of intervention, and continued to drop till 60 weeks (15 months), when it plateaued. Body weight also continued to decline till about 60 weeks, and then plateaued, reaching a new energy balance equilibrium during the 12-24mo period^61^. Thus, it is likely that the changes we observe in digalactosylation and the GlycAge during the 12-24mo period are a function of achieving and maintaining this new equilibrium in energy balance.

Glycosylation of total plasma protein, which includes several acute phase proteins, antithrombins, apolipoproteins, fibrinogens, and immunoglobulins^62^ showed a decrease in highly sialylated and galactosylated structures following CR intervention, with concomitant increases in asialyated glycans. In addition, there were decreased levels of highly branched (tri-and tetra-antennary) glycans in circulation following the intervention. While glycosylation is cell and tissue-type specific ^63^, plasma protein glycosylation patterns can provide overall insight into an individual’s immune and metabolic status and can differentiate healthy vs diseased states ^10,62^.

The reduced branched and highly sialylated glycans is a beneficial change in plasma protein glycome, since high branching and high sialylation of N-glycans is associated with increased inflammation and has been implicated in development of metabolic diseases ^64–66^. The increase in asialylation and agalactosylation, indicative of increased biological aging^14^ contrasts the above findings. Hence, certain plasma glycome beneficial changes from the 2y CR intervention were observed, while certain other biological aging related processes were not.

Following the 2-year CR intervention, complement C3 concentrations decreased in the cohort we studied. Complement C3 is a critical component of innate and adaptive immunological processes and originates from hepatic and occasionally, adipose tissue^67^. C3 concentrations are lower in naturally long-lived populations, e.g. centenarians, and higher in older individuals with metabolic syndrome and abdominal obesity^33,68^. Based on these prior reports, our results suggest that CR reductions of complement C3 are indicative of improvements in metabolic health and aging processes.

The link between the complement C3 glycome and aging/metabolic diseases are less understood. Here, CR reduced levels of Man9 and Man9Glc1 C3 glycoforms. Complement C3 glycoforms have been reported to be more “unprocessed,” (i.e. with more mannose and glucose sugars), in individuals with higher hemoglobin A1c and uncontrolled glycemia-associated vascular complications, compared to healthy counterparts ^30,34^. With 12 months of CR the Man9 and Man9Glc1 glycoforms were reduced, with reductions continued over 24 months for the Man9 glycans. The concurrent, 24 month increase in skeletal muscle expression of HSPA5 which encodes BiP, a UPR-associated chaperone, may indicate a means by which CR impacts ER stress through transcriptional and glycosylation processes to improve aging-related processes. Clearly, further investigation into this is necessary to improve our understanding.

## Limitations and future directions

This was a pilot study limited due to its small sample size consisting of 26 out of 188 completers of the CALERIE2 trial with an average of >10% CR attainment at 12 mo. Furthermore, we only studied the glycome of individuals in the CR group without comparison to the *ad libitum* group. Hence, future studies will aim to expand the cohort such that deeper insights can be obtained. We conclude that the effects of CR on these unique glycosylation biomarkers in this pilot study are worthy of future biochemical, genetic, cell, and animal studies evaluation and validation. Such analyses might reveal diagnostic and therapeutic glycan targets that are useful in monitoring and manipulating biological age. Glyco-engineered IgG proteins, which have desired glycosylation profiles (i.e. more sialylated, more galactosylated etc) are being investigated as anti-inflammatory and anti-auto-immune therapeutics^69^. These might also have anti-aging benefits, and our ability to glyco-engineer therapeutics rely on being able to better understand the effect of CR on the glycome.

## Data Availability

Data and select biological samples are available from the NIA AgingResearchBiobank upon request (https://agingresearchbiobank.nia.nih.gov/studies/calerie/)

