## Supplemental figures and tables for "A 2-year calorie restriction intervention reduces glycomic biological age biomarkers"

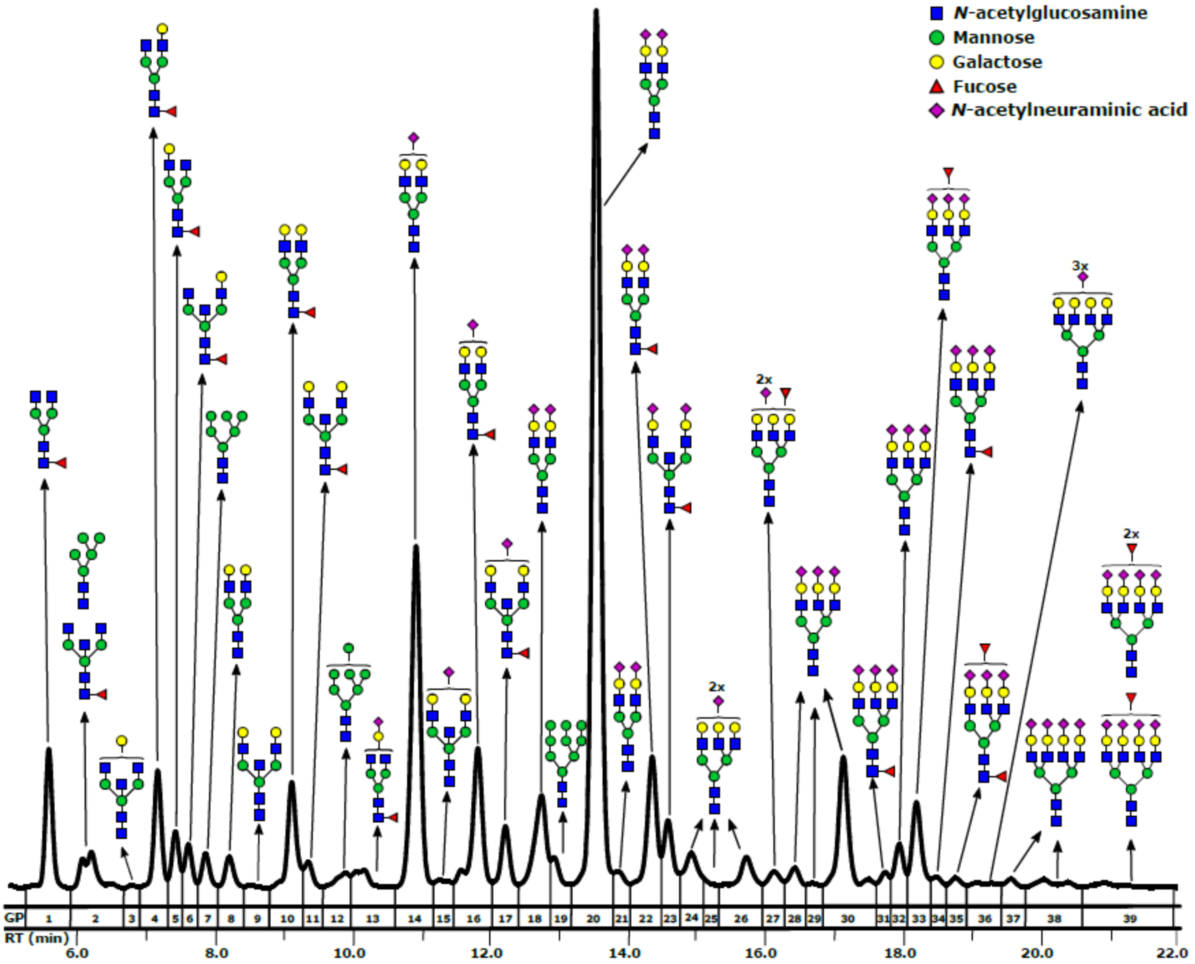


**Supplementary Figure 1** Representative HILIC-UPLC-FLR chromatogram of plasma protein N-glycome, with graphic representation of the most abundant glycan structures corresponding to each glycan peak (GP). RT – retention time.

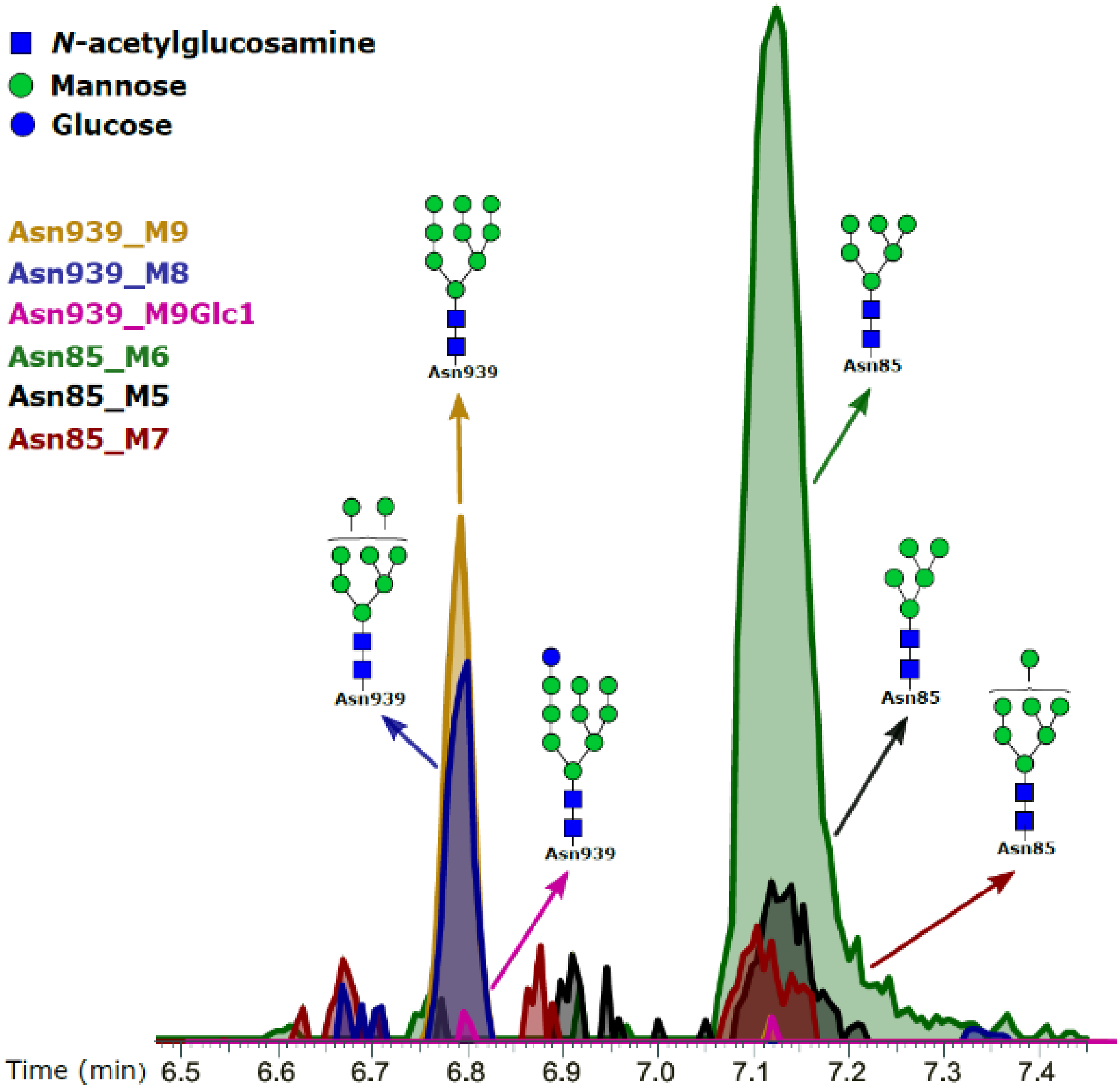


**Supplementary Figure 2** Representative EIC chromatogram of Complement C3 with graphic representation of the most abundant glycan structures in key.

**Supplementary Table 1: Plasma N-glycome derived traits using identified peaks (P1 – P39) in Supplemental Figure 1**

| Trait | Description | Calculation |
|---|---|---|
| LB | Low-branching | GP1 + GP2 + GP3 + GP4 + GP5 + GP6 + GP8 + GP9 + GP10 + GP11 + GP12 + GP13 + GP14 + GP15 + GP16 + GP17 + GP18 + GP20 + GP21 + GP22 + GP23 |
| HB | High-branching | GP24 + GP25 + GP26 + GP27 + GP28 + GP29 + GP30 + GP31 + GP32 + GP33 + GP34 + GP35 + GP36 + GP37 + GP38 + GP39 |
| S0 | Neutral/not sialylated | GP1 + GP2 + GP3 + GP4 + GP5 + GP6 + GP7 + GP8 + GP9 + GP10 + GP11 |
| S4 | Tetrasialylated | GP36 + GP37 + GP38 + GP39 |
| G0 | Agalactosylated | GP1 + GP2 |
| G4 | Tetragalactosylated | GP33 + GP34 + GP36 + GP37 + GP38 + GP39 |
| HM | High-mannose | GP2 + GP7 + GP19 |
| B | Bisecting N-acetylglucosamine | GP2 + GP3 + GP6 + GP9 + GP11 + GP12 + GP15 + GP17 + GP21 + GP23 |

**Supplemental Table 2: IgG N-glycome derived traits using identified peaks (P1 – P27) in manuscript figure 3A**

| Trait | Description | Calculation |
|---|---|---|
| G0 | agalactosylation | P14+P15+P18 |
| G1 | monogalactosylation | P16+P17+P19+P20+P21+P22+P23+P24 |
| G2 | digalactosylation | P25+P26+P27 |
| G | galactosylation (G1+G2) | P16+P17+P19+P20+P21+P22+P23+P24+P25+P26+P27 |
| S0 | asialylation | P14+P15+P16+P17+P18+P19+P20+P21+P22+P23+P24+P25+P26+P27 |
| F0 | afucosylation | P1+P2+P5+P6+P9+P10+P11+P14+P16+P17+P19+P20+P25 |
| B | bisecting GlcNAc | P2+P4+P11+P13+P14+P18+P19+P20+P23+P24+P25+P27 |
| B0 | absence of bisecting GlcNAc | P1+P3+P5+P6+P7+P8+P9+P10+P12+P15+P16+P17+P21+P22+P26 |

Supplemental Table 3: Summary of all outcomes

| | | | | | | ANCOVA model* | | |
|---|---|---|---|---|---|---|---|---|
| **Parameter** | **Month 0** | **Month 12** | **Month 24** | **Δ 12mo** | **Δ 24mo** | **Month** | **Sex** | **Age** |
| GlycAge (y) | 38.3 ± 7.8 | 38.9 ± 7.5 | 37.7 ± 8.1 | 0.60 ± 0.26 | -0.54 ± 0.25 | **0.032** | 0.537 | - |
| **IgG** | | | | | | | | |
| Agalactosylation | 19.9 ± 5.5 | 19.9 ± 5.3 | 19.3 ± 5.4 | -0.01 ± 0.17 | -0.56 ± 0.06 | 0.073 | 0.514 | 0.786 |
| Digalactosylation | 21.5 ± 4.3 | 21.2 ± 4.1 | 21.9 ± 4.3 | -0.22 ± 0.18 | 0.43 ± 0.05 | **0.025** | 0.298 | 0.798 |
| Galactosylation (G1+G2) | 59.5 ± 3.6 | 59.7 ± 3.7 | 60.1 ± 3.7 | 0.24 ± 0.09 | 0.61 ± 0.10 | **0.036** | 0.396 | 0.875 |
| Asialylation | 79.4 ± 2.7 | 79.6 ± 2.4 | 79.4 ± 2.7 | 0.23 ± 0.39 | 0.05 ± 0.07 | 0.372 | 0.274 | 0.219 |
| Afucosylation | 4.5 ± 0.9 | 4.6 ± 1.0 | 4.5 ± 1.0 | 0.10 ± 0.10 | 0.02 ± 0.06 | 0.21 | 0.996 | 0.134 |
| Bisecting GlcNAc | 16.9 ± 2.1 | 17.4 ± 2.3 | 17.3 ± 2.1 | 0.42 ± 0.15 | 0.31 ± 0.05 | **0.0004** | 0.299 | 0.231 |
| **Plasma** | | | | | | | | |
| Asialylation | 22.9 ± 4.3 | 23.6 ± 4.1 | 24.4 ± 3.7 | 0.65 ± 0.17 | 1.49 ± 0.62 | **0.0012** | 0.349 | 0.496 |
| Tetrasialylation | 2.6 ± 0.5 | 2.5 ± 0.5 | 2.3 ± 0.5 | -0.03 ± 0.02 | -0.21 ± 0.04 | **0.0148** | 0.297 | 0.114 |
| Agalactosylation | 6.1 ± 1.8 | 6.3 ± 1.8 | 6.4 ± 1.6 | 0.17 ± 0.01 | 0.23 ± 0.19 | **0.0411** | 0.2207 | 0.2143 |
| Tetragalactosylation | 5.3 ± 1.3 | 5.3 ± 1.2 | 4.9 ± 1.1 | 0.00 ± 0.13 | -0.38 ± 0.22 | **0.0187** | 0.0771 | 0.0925 |
| Oligomannose | 4.2 ± 0.4 | 4.2 ± 0.4 | 4.3 ± 0.4 | -0.03 ± 0.04 | 0.08 ± 0.02 | **0.0368** | 0.6087 | 0.864 |
| Bisecting GlcNAc | 10.2 ± 1.2 | 10.6 ± 1.3 | 11.0 ± 1.1 | 0.34 ± 0.08 | 0.76 ± 0.05 | **<0.0001** | 0.917 | 0.5951 |
| High-branched | 17.5 ± 2.5 | 17.0 ± 2.5 | 16.2 ± 2.2 | -0.51 ± 0.04 | -1.27 ± 0.28 | **0.002** | 0.5501 | 0.6803 |
| **Complement C3** | | | | | | | | |
| Total Complement C3 (g/L) | 1.19 ± 0.24 | 1.06 ± 0.22 | 1.04 ± 0.16 | -0.13 ± 0.02 | -0.15 ± 0.08 | **<0.0001** | 0.138 | 0.103 |
| C3_Asn_85_N2H5 | 0.133 ± 0.013 | 0.132 ± 0.017 | 0.125 ± 0.013 | -0.001 ± 0.005 | -0.008 ± 0.000 | **0.0028** | 0.8751 | 0.2357 |
| C3_Asn_85_N2H6 | 0.790 ± 0.017 | 0.803 ± 0.016 | 0.800 ± 0.017 | 0.013 ± 0.001 | 0.010 ± 0.000 | **<0.0001** | 0.9789 | 0.7609 |
| C3_Asn_85_N2H7 | 0.077 ± 0.016 | 0.065 ± 0.016 | 0.075 ± 0.021 | -0.012 ± 0.000 | -0.002 ± 0.005 | **<0.0001** | 0.7231 | 0.363 |
| C3_Asn_939_N2H8 | 0.365 ± 0.035 | 0.384 ± 0.037 | 0.379 ± 0.036 | 0.019 ± 0.002 | 0.014 ± 0.002 | **<0.0001** | 0.2202 | 0.1343 |
| C3_Asn_939_N2H9 | 0.590 ± 0.031 | 0.576 ± 0.034 | 0.580 ± 0.031 | -0.015 ± 0.003 | -0.011 ± 0.000 | **<0.0001** | 0.3188 | 0.2771 |
| C3_Asn_939_N2H10 | 0.044 ± 0.010 | 0.040 ± 0.007 | 0.041 ± 0.009 | -0.004 ± 0.003 | -0.003 ± 0.000 | **0.0152** | 0.3809 | 0.3709 |

* BL used as covariate along with BL x month interaction
